# Gate-Before-Generate: A Dual-Layer Architecture for Output-Presence Routing in Chest X-ray Report Generation

**DOI:** 10.64898/2026.08.11.26360223

**Authors:** Tung-Chu Bai, Sheng-Cheng Yeh

## Abstract

**Background:** CXR report generation may require a vision-language model (VLM) to produce both textual findings and spatial bounding boxes. Generative 4B–7B VLMs can emit non-empty outputs on normal images and empty outputs on abnormal images, motivating explicit structural routing.

**Objective:** To evaluate whether a hard inference-time gate before a probabilistic VLM changes output-presence performance and to identify the mechanisms underlying paired STRUCT outcomes.

**Methods:** We evaluated CXRxVLM v2, combining a frozen microsoft/rad-dino ViT-B/14 encoder with a 768*→*1 logistic probe (threshold 0.0557) and google/medgemma-4b-it with the pamessina/medgemma-4b-it-cure LoRA adapter. A seed=42 stratified cohort of 500 VinDr-CXR train-pool images (250 NORMAL, 250 ABNORMAL) was compared with Lingshu-7B A_baseline and D_fewshot configurations. Exact paired McNemar tests and stratum-level output-presence analyses were prespecified for the primary configurations; MedGemma 1.5 SigLIP was exploratory.

**Results:** CURE achieved STRUCT = 78.0% (390/500; Wilson 95% CI 74.2–81.4), versus 73.8% for Lingshu A_baseline and 74.2% for D_fewshot. Pairwise *p*-values were 0.0778, 0.1042, and 0.8642. The paired decomposition showed CURE ABNORMAL non-empty-output advantage of +13.6 percentage points versus Lingshu A (*p* = 0.0012; +14.0 points versus D, *p* = 0.0007), while Lingshu had higher NORMAL empty-output rates (+5.2 to +6.4 points; *p* = 0.0106 and *p* = 0.0004). The full pipeline used 8.87 GB VRAM and 4.92 s/image mean latency; 53% of records used a 25.7 ms warm gate-negative path after model loading.

**Conclusions:** Equivalent overall STRUCT scores concealed two mechanistically different output regimes: CURE favored ABNORMAL non-empty outputs, whereas Lingshu favored NORMAL empty outputs. This paired decomposition, rather than the aggregate score alone, characterizes how hard-gated and probabilistic systems route output presence.

## 1 Introduction

### 1.1 Clinical and computational motivation

Chest X-ray (CXR) is the most frequently performed radiographic examination worldwide, with more than two billion studies conducted annually [1]. Some clinical and research workflows may benefit from both a *textual* description of findings (e.g., “cardiomegaly, mild pulmonary edema”) and a *spatial* indication of a lesion’s location on the original image. Producing both simultaneously — known as *grounded report generation* — is an important research problem in medical-image understanding. The emergence of 4B–7B vision-language models (VLMs) [2–5] has made automatic CXR report drafting technically feasible, but three systematic limitations remain unsolved at inference time:

1. **Hallucination.** VLMs can produce clinically problematic outputs: an abnormal image may be reported as “no finding”, or a normal image may receive a spurious bbox. CURE reports an author-evaluated hallucination rate of 8.78% under the authors’ original definition and evaluation protocol; that value is not directly comparable with the STRUCT endpoint reported here [5].
2. **Localization precision.** Clinical interpretation would require reliable lesion-level localization; this study evaluates the binary presence of at least one bbox, not IoU- or label-level localization correctness.
3. **Deployment cost.** Full VLM inference requires substantially more VRAM than a lightweight gate; the CURE stack in this study uses 8.87 GB in 4-bit quantization, whereas the gate-only path uses 0.35 GB.

Prior work addresses these limitations through either *training-time* regularization (SFT with normal-image-weighted sampling, RL with hallucination penalties) or *post-hoc* filtering (confidence thresholds, language-model judges). Both are intrinsically *probabilistic*: they can reduce but cannot structurally eliminate non-empty outputs on normal images, because a generative decoder still has a non-zero probability of emitting a token for any input.

### 1.2 The structural-vs-probabilistic gap

An inference-time hard gate can provide a zero-bbox output guarantee for the subset of images it labels NORMAL: a binary classifier short-circuits those images before the VLM is invoked [6, 7]. We use this distinction to separate an explicit structural decision from probabilistic report generation, and evaluate both layers end to end with paired McNemar tests and discordant-pair decomposition.

### 1.3 Research questions

This study answers three questions on a 500-image, seed=42 stratified VinDr-CXR subset:

1. **Ceiling.** Does a dual-layer Gate-Before-Generate architecture (CURE configuration) exceed the STRUCT score of the two primary Lingshu evaluation configurations by a statistically significant margin? The MedGemma 1.5 arm is considered separately and is not included in the primary paired analysis.
2. **Decomposition.** If the systems differ, how do the NORMAL empty-output and ABNORMAL non-empty-output rates differ across the tested configurations?
3. **Prompt robustness.** Does prompt tuning of Lingshu-7B (A_baseline *→* D_fewshot) that was significant on a 50-image pilot remain significant on the 500-image evaluation, or is it a small-sample artefact?

### 1.4 Contributions

- **ARCHITECTURAL:** A hard structural gate provides a zero-bbox guarantee for the gate-negative subset, categorically distinguishing it from probabilistic mitigations such as SFT, RL, and threshold filters.
- **EMPIRICAL:** A paired McNemar decomposition on a 500-image cohort reveals two mechanistically different output regimes despite equivalent overall STRUCT, a result invisible without stratum-level paired analysis.
- **DEPLOYMENT:** The full dual-layer pipeline runs in 8.87 GB VRAM at 4.92 s/image mean latency, with 53% of images short-circuited to a 25.7 ms warm path through the 769-parameter gate.

## 2 Related Work

### 2.1 Evolution of CXR vision-language models

CXR VLM research can be divided into three phases. **Phase 1 (2017–2020)** comprised task-specific CNNs (CheXNet [1], DenseNet-121 baselines) for classification only. **Phase 2 (2020–2024)** introduced large-scale supervised pre-training on CheXpert [2] and MIMIC-CXR [3], scaling model capacity but not addressing grounded reporting. **Phase 3 (2025–2026)** combines self-supervised vision encoders with 4B–7B VLMs; CXRxVLM v2 belongs to this phase.

### 2.2 2025–2026 CXR VLM landscape

Table 1 summarizes systems relevant to this study and their status in our evaluation. First, the compared probabilistic systems do not provide a structural zero-bbox guarantee before generation. Second, AnatomiX [8] and CXRMate-2 [9] pursue supervised fine-tuning or reinforcement learning on labeled CXR data and therefore fall outside the no-additional-task-specific-VLM-training constraint adopted here. Third, larger Lingshu variants imply substantially higher hardware requirements than the 7B configuration evaluated in this study.

**Table 1.** 2025–2026 CXR vision-language model comparison (relevant systems).

| Method | Type | Year | Ground. | CXR STRUCT in this study |
| --- | --- | --- | --- | --- |
| CheXNet [1] | CNN (DenseNet-121) | 2017 | No | N/A (classification only) |
| MedGemma 1.5-4B [10] | VLM (SigLIP 1152-d) | 2026 | Partial | Exploratory; no paired artifact deposited |
| Lingshu-7B [4] | VLM (Qwen2.5-VL) | 2026 | Yes | 73.8–74.2% (two prompts, this study) |
| Lingshu-I-8B [4] | VLM (InternVL3) | 2026 | Yes | Not tested (vLLM Windows path fail) |
| CURE [5] | LoRA on MedGemma-4B | 2026 | Yes | N/A here; 8.78% under the published protocol, not directly comparable to STRUCT |
| **CXRxVLM v2** | **Dual-layer gate + VLM** | **2026** | **Yes** | **78.0% [74.2–81.4]** |
| Anatomix [8] | SFT CXR VLM | 2026 | Yes | Excluded (requires SFT) |
| CXRMate-2 [9] | RL-tuned CXR VLM | 2026 | Yes | Excluded (requires RL) |

### 2.3 Why an inference-time hard gate is structurally different

Probabilistic mitigations (SFT, RL, contrastive regularization) optimize an expected reward; they do not impose a hard zero-bbox decision before generation. Related work on visual alignment and perception-centric process rewards likewise operates within probabilistic inference [11, 12]. The CXRxVLM v2 architecture adds a *binary structural decision* at the gate, which — by construction — emits zero bboxes on any image the gate labels as NORMAL.

## 3 Methods

### 3.1 System architecture

The dual-layer Gate-Before-Generate architecture is shown in Fig. 1. Every input CXR image is first encoded by the gate; only images the gate classifies as ABNORMAL are forwarded to the VLM.

**Figure 1.**
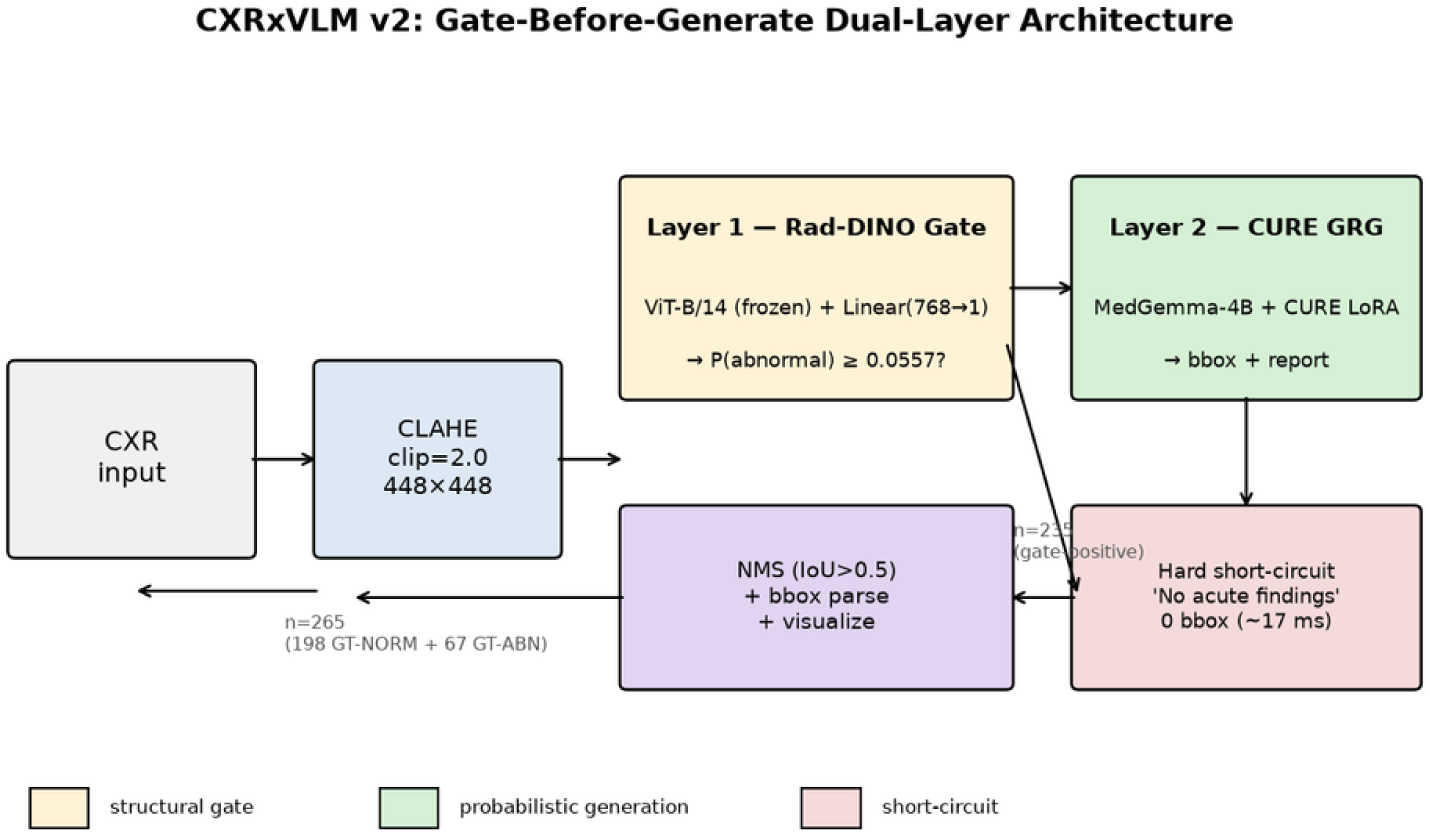
Dual-layer Gate-Before-Generate architecture. Gate-negative images terminate with a research-only zero-bbox output; gate-positive images proceed to probabilistic CURE generation. The figure depicts output routing, not localization or clinical correctness.

**Figure 2.**
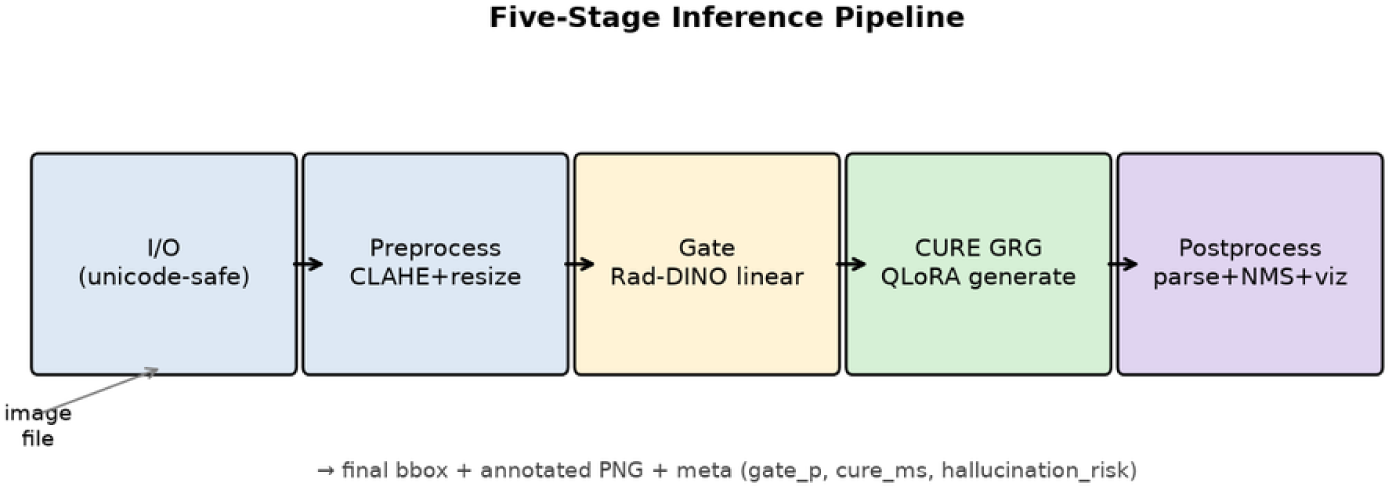
Evaluation and inference workflow for the five-stage pipeline. The 500-image shared-pool cohort was processed through gate evaluation, optional CURE generation, and output-presence scoring.

The two inference paths are summarized in Fig. S1. The gate-only path returns a structural zero-bbox result for images below the operating threshold; gate-passing images continue through CURE generation and post-processing.

**Layer 1 — Rad-DINO linear-probe gate (structural).** The frozen microsoft/rad-dino ViT-B/14 encoder produces a 768-dimensional embedding using the evaluated mean-pooling implementation; a single logistic-regression linear probe (768*→*1) followed by sigmoid yields *P*(abnormal). The operating threshold is *τ* = 0.0557, selected during gate development to satisfy the target dataset-label operating point. Images with *P*(abnormal) *< τ* are short-circuited with a research-only “below gate threshold” status and zero bboxes; this is not a normal finding, negative examination, exclusion decision, or clinical screening result. These outputs are research-only and should not be used as a substitute for physician review or clinical decision-making. A separate gate-only microbenchmark reported approximately 17 ms per image on an RTX 5090; this excludes VLM inference and evaluator setup overhead.

**Layer 2 — CURE GRG (probabilistic).** Images passing the gate are preprocessed (Section 3.2) and forwarded to google/medgemma-4b-it quantized to NF4 with the pamessina/medgemma-4b-it-cure LoRA adapter (rank 16, alpha 16, all-linear) [13, 14]. The model is prompted with “Generate a grounded report.” and produces a free-text report plus CURE-style [cx, cy, w, h] token-bboxes. The observed latency distribution is reported from the 500-image evaluation run in Section 4.10.

**Post-processing.** The cxcywh tokens are converted to xyxy pixel coordinates, non-maximum suppression (NMS, IoU *>* 0.5) is applied, and the resulting bboxes are visualized on the original image.

### 3.2 Pre-processing

We observed that the choice of CLAHE clipLimit was associated with a 4-percentage-point STRUCT difference in the 50-image pilot: clipLimit = 2.0 with tileGridSize = (8, 8) (corrected from the original 3.0 in CURE’s training pipeline) yielded STRUCT 88%, compared with 84% at clipLimit = 3.0. The image is then resized to 448*×*448 and normalized with ImageNet statistics.

### 3.3 Gate training

The gate is the only trainable component. We froze the Rad-DINO encoder, extracted 768-dimensional patch-token mean-pooled features from the 12,000-image VinDr-CXR train.csv/images/train pool (8,489 NORMAL, 3,511 ABNORMAL; 70.7%/29.3%), and fit a scikit-learn logistic-regression probe with *C* = 1.0, max_iter = 2000, and positive-class weight 6.0. Model fitting used an 80/20 stratified split with random seed 42. The exact image IDs assigned to the historical training and calibration partitions were not preserved, so overlap between those partitions and the 500-image evaluation cohort cannot be determined retrospectively. The checkpoint metadata records the mean-pooling configuration and calibration summary. Validation AUC = 0.9826 (rounded from checkpoint metadata); the threshold was selected during gate development for the target dataset-label operating point.

### 3.4 Evaluation methodology

A 500-image subset was sampled from the VinDr-CXR ‘train.csv‘/‘images/train‘ pool (250 NORMAL, 250 ABNORMAL) [15] with np.random.RandomState(42) stratified by the official label. All three primary JSONL artifacts contain the same 500 image IDs. Because the historical gate feature cache did not preserve image IDs or split assignments, exact train/evaluation disjointness cannot be proven retrospectively; we therefore treat this as a shared-pool retrospective benchmark rather than an independent test evaluation and do not present the gate AUC as independent validation evidence. For each system we computed two output criteria per image: a NORMAL empty-output rate (no bbox) and an ABNORMAL non-empty-output rate (*≥* 1 bbox). STRUCT is defined as (*N*_NORMAL empty_ + *N*_ABN non-empty_) / 500. For every paired difference below, positive values mean higher correct-output presence for the first-named system. These are output-presence proxies, not measures of localization, label correctness, report correctness, or clinical sensitivity.

### 3.5 Rationale for the STRUCT endpoint

STRUCT is deliberately a binary, stratum-level output-presence proxy: it is a first-pass architectural sanity check that separates correct pipeline routing from localization agreement. A grounded VLM that emits bboxes on normal images, or emits an empty output on abnormal images, fails this routing test regardless of the quality of any localization it produces. STRUCT therefore exposes routing failures with paired statistical power. Downstream IoU-based localization evaluation and clinical adjudication are future work.

The primary STRUCT comparisons have *n*_disc_ = 34–129; the NORMAL empty-output comparisons have *n*_disc_ = 5–23 and the ABNORMAL non-empty-output comparisons *n*_disc_ = 29–106. Each is reported with its exact reliability status: VALID for *n*_disc_ *≥* 25, LIMITED for 10 *≤ n*_disc_ *<* 25, and UNRELIABLE for *n*_disc_ *<* 10. The primary endpoint is final output presence; the stratum analyses are secondary descriptive comparisons. We report observed differences without interpreting non-significance as equivalence.

### 3.6 Hardware and software

All experiments were performed on a single workstation with one NVIDIA RTX 5090 (Blackwell, SM_120, 32 GB), an Intel Core Ultra 9 285 CPU, and 64 GB DDR5 RAM. The CURE stack ran in NF4 4-bit quantization (peft 0.19.1, bitsandbytes, transformers 5.12.1, torch 2.11+cu128). Lingshu-7B ran in fp16 via the HuggingFace transformers generate pipeline. Total VRAM at peak: 8.87 GB.

## 4 Results

The main-figure sequence is organized as follows: architecture (Fig. effig:arch), evaluation design and cohort (Table eftab:protocol), sample convergence (Fig. effig:converge), primary STRUCT comparison (Fig. effig:struct), stratum trade-off (Fig. effig:tradeoff), and failure taxonomy (Fig. effig:fail). Less central implementation and exploratory material is retained in the Supplementary Materials.

### 4.1 Sample size and evaluation scope

We report the 50- and 200-image values as historical, non-reconstructed development estimates; the corresponding immutable per-image artifacts and analysis inputs are not part of the current reproducibility package. We compare them descriptively with the selected 500-image estimate. The Wilson 95% interval narrows as the subset grows: it spans approximately [*−*13.0, +7.5] percentage points around the 88.0% estimate at *n* = 50, [*−*7.0, +6.2] around 79.5% at *n* = 200, and [*−*3.8, +3.4] around 78.0% at *n* = 500. This narrowing is descriptive and does not establish statistical saturation, external validity, or that a larger evaluation is unnecessary. We report 78.0% [74.2–81.4] as the estimate for this selected shared-pool cohort.

### 4.2 Three-system STRUCT comparison

The three primary evaluation configurations produce the following point estimates on the 500-image evaluation: CURE configuration 78.0% (390/500), Lingshu-7B A_baseline 73.8% (369/500), Lingshu-7B D_fewshot 74.2% (371/500). For CURE, this final output-presence score combines 231/250 empty-output NORMAL images and 159/250 non-empty-output ABNORMAL images; the corresponding gate-only binary output-routing agreement score is 76.2% (198/250 gate-negative NORMAL and 183/250 gate-positive ABNORMAL). The three pairwise STRUCT differences are shown in Fig. 4 and Table 3.

**Figure 3.**
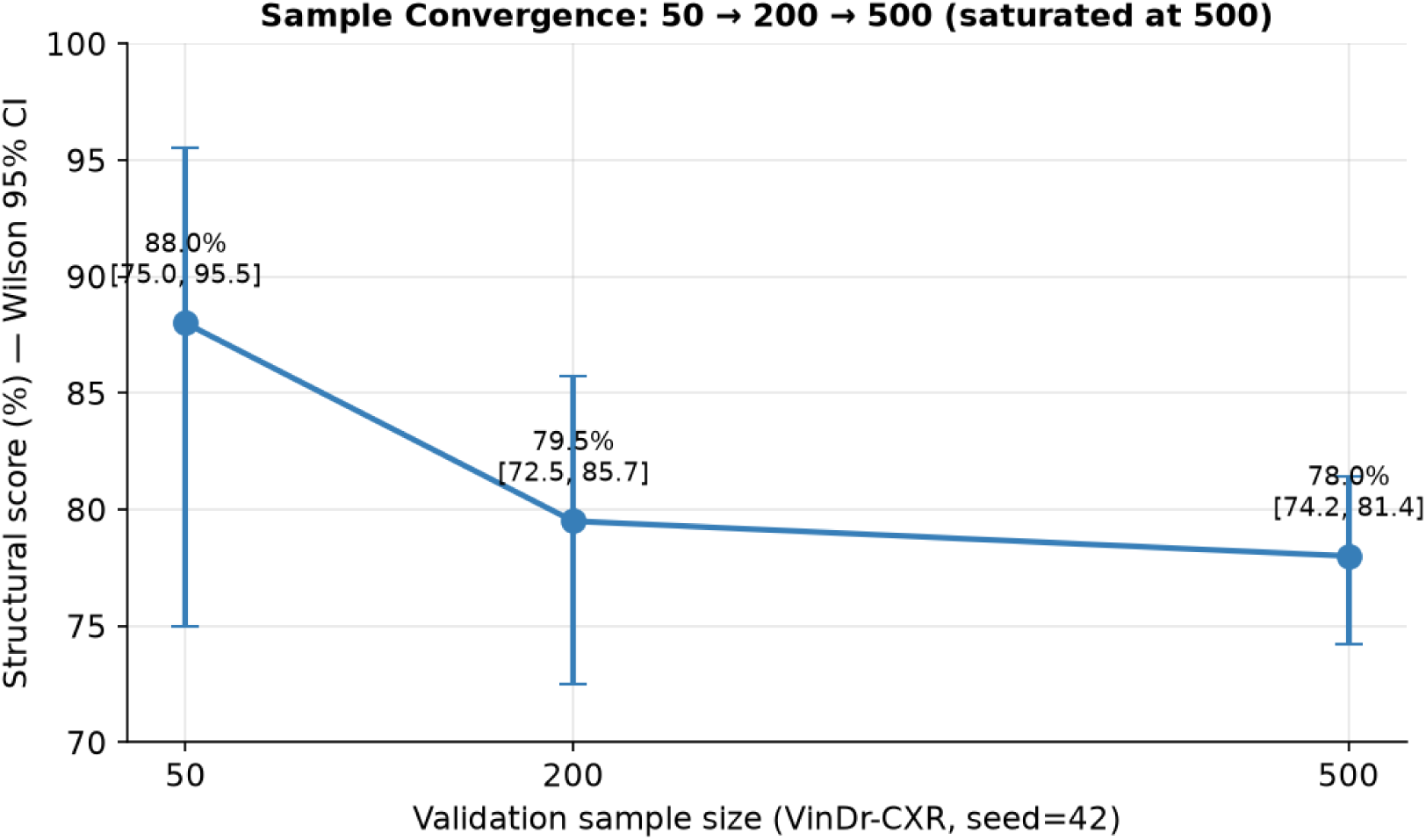
Sample convergence 50 *→* 200 *→* 500 (CURE configuration, seed=42).

**Figure 4.**
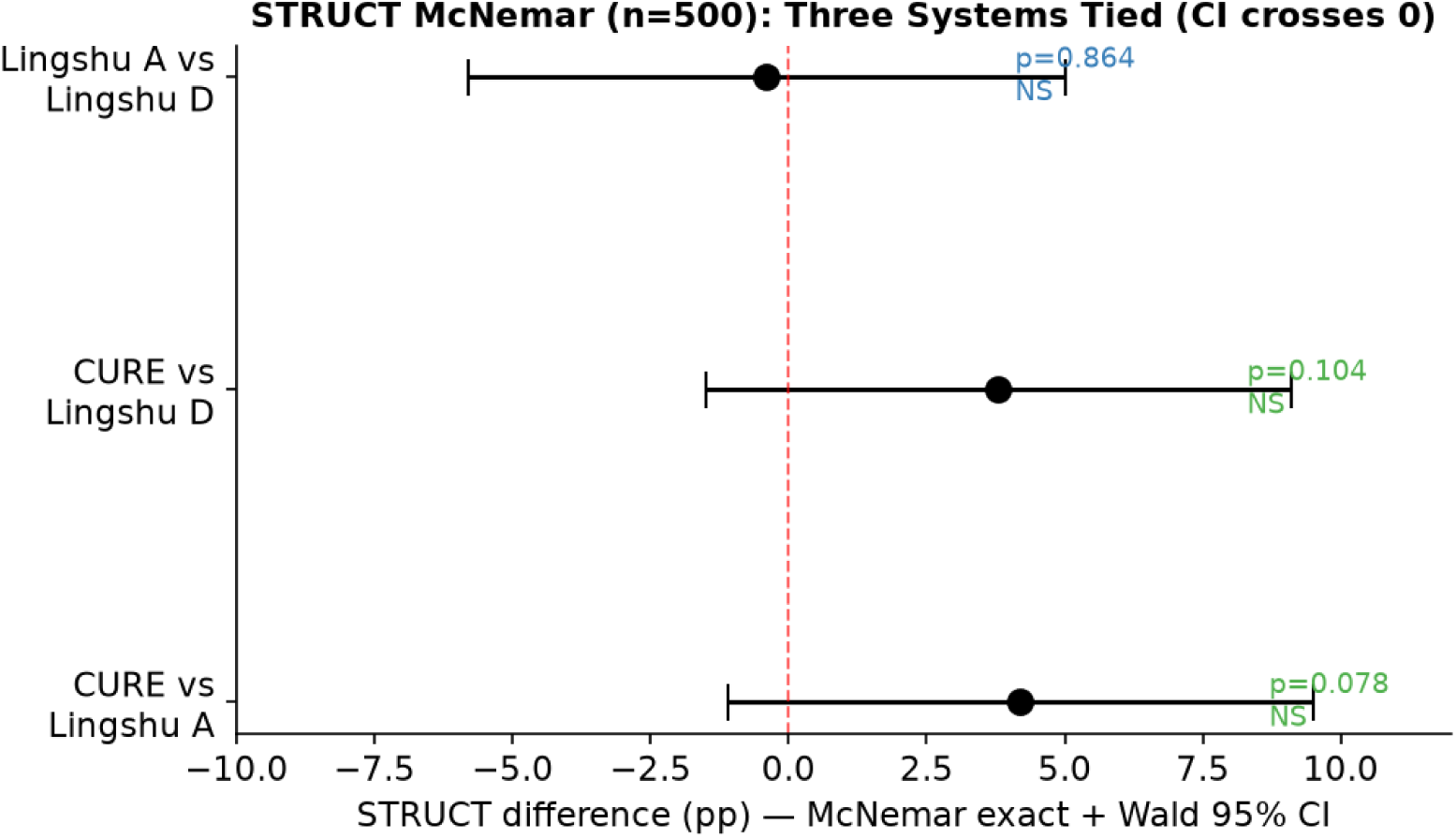
NORMAL empty-output rate and ABNORMAL non-empty-output rate: paired secondary comparisons (*n* = 250 per stratum).

**Table 2.** Statistical comparison protocol.

| Quantity | Definition |
| --- | --- |
| discordant pair $(b, c)$ | $b = \#\{A = 1, B = 0\}$; $c = \#\{A = 0, B = 1\}$ on the same 500 images |
| $n_{\text{disc}}$ | $b + c$ |
| McNemar exact $p$ | $2 \min\{\Pr[B \leq \min(b, c)], \Pr[B \geq \max(b, c)]\}$, $B \sim \text{Binom}(b + c, 0.5)$ |
| Paired 95% CI | Discordant-pair Wald approximation for the correct-output difference; descriptive uncertainty alongside exact McNemar $p$ |
| Wilson 95% CI (single) | Wilson score interval for individual proportions |
| $n_{\text{disc}}$ reliability | VALID $\geq 25$ / LIMITED 10–24 / UNRELIABLE $< 10$ |

**Table 3.** STRUCT McNemar paired tests (*n* = 500). Differences are first-named system minus second-named system; positive values favor the first-named system.

| Comparison | $n_{\text{disc}}$ | $p_{\text{exact}}$ | 95% CI on diff (pp) | Conclusion |
| --- | --- | --- | --- | --- |
| CURE vs Lingshu A | 129 | 0.0778 | +4.2 [−0.19, +8.59] | NS (VALID; CI crosses 0) |
| CURE vs Lingshu D | 123 | 0.1042 | +3.8 [−0.50, +8.10] | NS (VALID; CI crosses 0) |
| Lingshu A vs D | 34 | 0.8642 | −0.4 [−2.68, +1.88] | NS (VALID; D_fewshot no real improvement) |

**Table 4.** NORMAL empty-output paired comparisons (*n* = 250 NORMAL subset). Differences are first-named minus second-named correct-output rates; positive values favor the first-named system.

| Comparison | $n_{\text{disc}}$ | $p_{\text{exact}}$ | 95% CI on diff (pp) | Conclusion |
| --- | --- | --- | --- | --- |
| CURE vs Lingshu A | 23 | 0.0106 | -5.20 [-8.30, -2.10] | Lingshu significantly higher (LIMITED) |
| CURE vs Lingshu D | 20 | 0.0004 | -6.40 [-8.50, -4.30] | Lingshu significantly higher (LIMITED) |
| Lingshu A vs D | 5 | 0.3750 | -1.20 [-2.60, +0.20] | NS (UNRELIABLE: $n_{\text{disc}} < 10$) |

**Table 5.** ABNORMAL non-empty-output paired comparisons (*n* = 250 ABNORMAL subset). Differences are first-named minus second-named correct-output rates; positive values favor the first-named system.

| Comparison | $n_{\text{disc}}$ | $p_{\text{exact}}$ | 95% CI on diff (pp) | Conclusion |
| --- | --- | --- | --- | --- |
| CURE vs Lingshu A | 106 | 0.0012 | +13.60 [+5.95, +21.25] | CURE significantly higher (VALID) |
| CURE vs Lingshu D | 103 | 0.0007 | +14.00 [+6.52, +21.48] | CURE significantly higher (VALID) |
| Lingshu A vs D | 29 | 1.0000 | +0.40 [-3.82, +4.62] | NS (VALID, D_fewshot no real improvement) |

### 4.3 NORMAL empty-output and ABNORMAL non-empty-output decomposition

The point estimates split when STRUCT is decomposed into the NORMAL empty-output rate and ABNORMAL non-empty-output rate on the 250-image strata: CURE configuration 92.4% / 63.6%, Lingshu-7B A 97.6% / 50.0%, and Lingshu-7B D 98.8% / 49.6%.

The decomposition shows a trade-off in output-presence rates on this cohort: Lingshu-7B has higher NORMAL empty-output rates, whereas CURE has higher ABNORMAL non-empty-output rates. The trade-off is visualized in Fig. 5.

**Figure 5.**
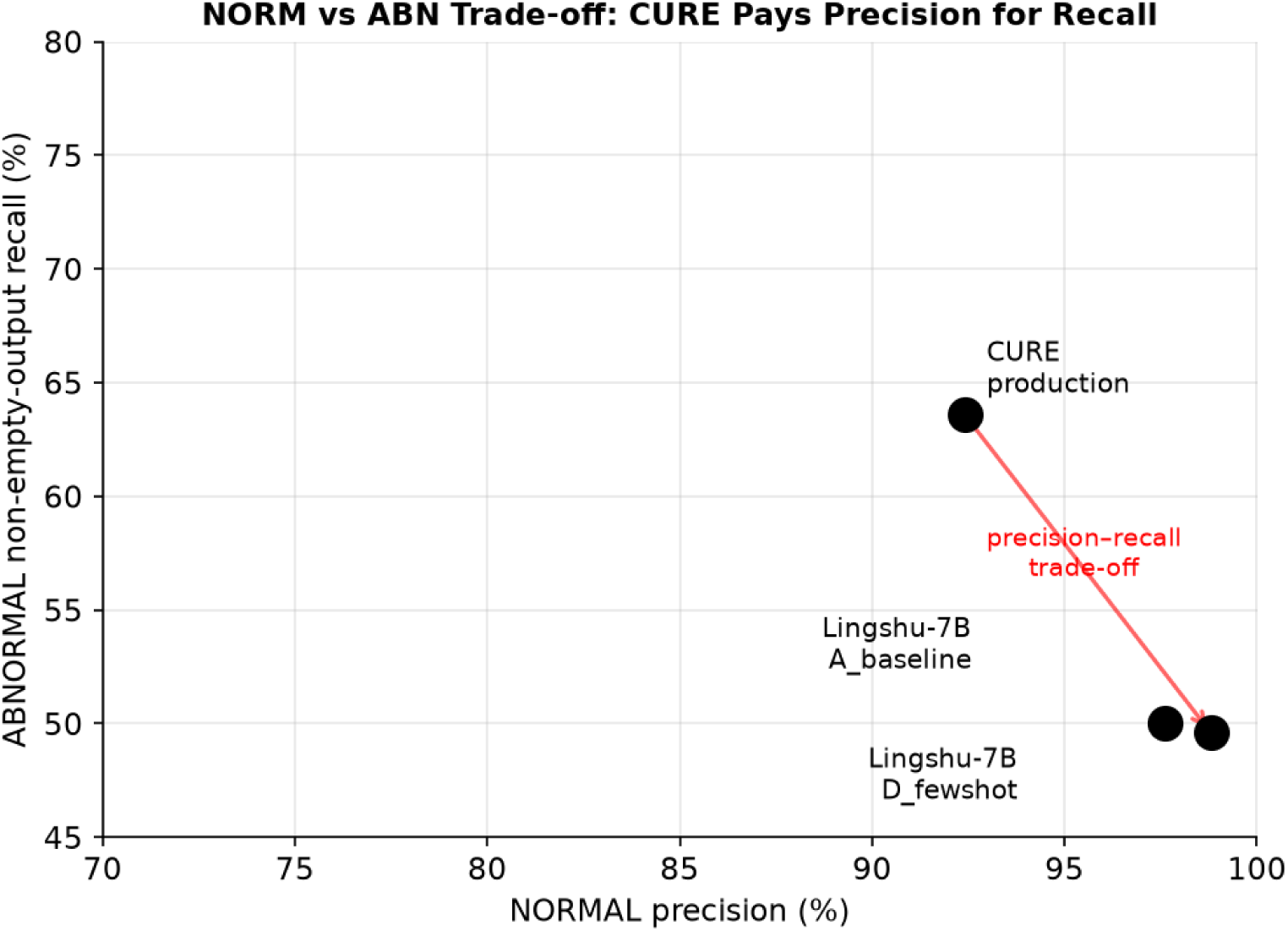
NORMAL empty-output rate versus ABNORMAL non-empty-output rate on the shared evaluation cohort.

**Figure 6.**
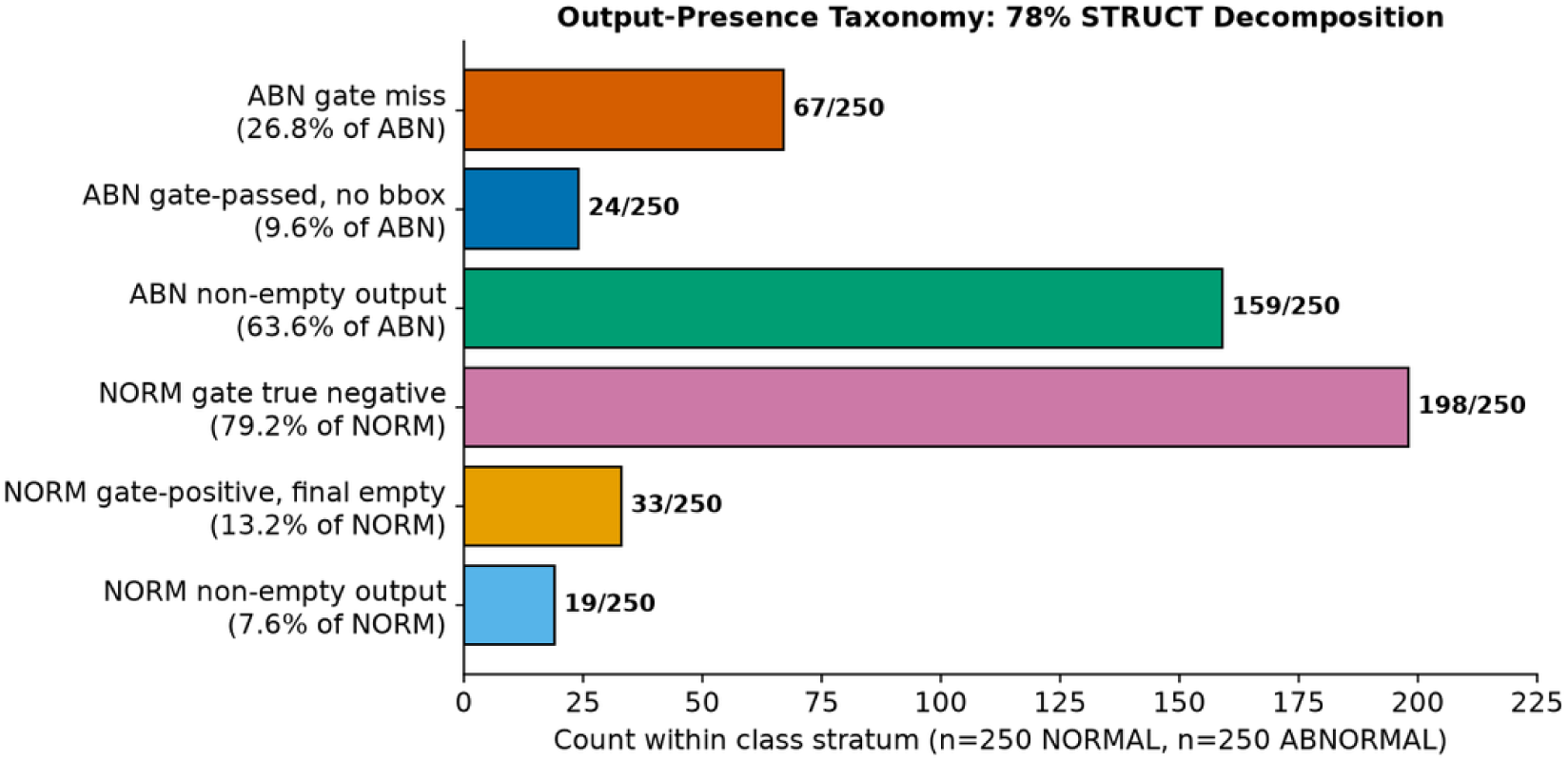
Output-presence taxonomy by class stratum (*n* = 250 NORMAL, *n* = 250 ABNORMAL); categories do not assert semantic hallucination or localization correctness.

### 4.4 MedGemma 1.5 SigLIP gate (exploratory arm)

MedGemma 1.5 was examined in a separate exploratory gate experiment because its cached-feature validation behavior differed from the Rad-DINO production gate. The full-pipeline per-image records for that arm are not included in the released evaluation artifact; consequently, we do not report a reproducible STRUCT estimate or perform a paired inferential comparison for MedGemma 1.5. The arm is retained only as a documented exploratory direction and is excluded from all primary tables and McNemar analyses.

### 4.5 Failure-mode taxonomy

The 500-image CURE configuration result decomposes into six mutually exclusive outcomes. Among the 250 ABNORMAL images, 67 (26.8%) are missed by the gate and never reach the VLM, 24 (9.6%) pass the gate but produce no bbox, and 159 (63.6%) produce at least one bbox under the STRUCT output-presence criterion. Among the 250 NORMAL images, 198 (79.2%) are short-circuited by the gate, 33 (13.2%) are gate-positive but produce zero bboxes through CURE, and 19 (7.6%) produce a non-empty output. The final STRUCT score is 78.0% because it counts all 231 NORMAL empty-output cases, including 33 gate-positive CURE rescues, plus 159 ABNORMAL non-empty-output cases.

### 4.6 CLAHE pre-processing ablation

The single largest pre-processing effect identified was the CLAHE clipLimit. Fig. S10 shows side-by-side CLAHE-clip=3.0 (legacy, the CURE training default) and clip=2.0 (this work’s correction) on a multi-focal abnormal image. The clip=2.0 image reveals a second lesion in the right-lower zone that the clip=3.0 image washes out; on the 50-image pilot this corresponds to a 4-percentage-point STRUCT improvement.

### 4.7 Real case studies

Five representative real-image cases are shown in Figs. S3–S7. Each panel is a ground-truth reference: the original VinDr-CXR validation image with the official annotations (green), without model predictions overlaid. The cases span clean NORMAL anatomy, multi-class abnormal findings, the smallest annotated lesion in the source set, a diffuse abnormal pattern, and a multi-annotation abnormal example.

### 4.8 SFT text-quality failure mode

A separate failure mode — text-quality collapse under supervised fine-tuning — is illustrated in Fig. S12. The pretrained CURE-adapter configuration emits anatomically specific labels (e.g., “Cardiomegaly. Dorsal scoliosis.

Metallic sutures in the midline.”); the LoRA-SFT model collapses to a generic “finding” pattern, which appears less anatomically specific in the inspected examples. We documented this negative result in a separate analysis; SFT was therefore not adopted for production.

### 4.9 Hallucination-risk flag distribution

The F-17 hallucination-risk flag is a runtime check that triggers recommendation:manual_review for any of four implemented conditions: (1) a gate-positive image with zero bboxes; (2) empty VLM output; (3) no recognized bbox label in the implementation’s known-label allowlist; and (4) gate probability within *±*0.1 of the operating threshold. Because the runtime returns before F-17 assessment on gate-negative images, the audit population was the 183 gate-positive images within the ground-truth ABNORMAL stratum; the 67 gate-missed abnormalities were not F-17-checked. The retained evaluation JSONL exposes only a boolean hallucination_risk field, yielding 24 flagged ABNORMAL records (24/250 of all ABNORMAL images and 24/183 of the gate-positive audit population). It does not retain per-condition flags or the raw fields needed to independently reconstruct the other three condition counts, so those counts are not reported. Fig. S9 reports the retained evidence only and should not be interpreted as the total runtime flag count. A non-medical-vocabulary check remains a documented future extension rather than an implemented trigger.

### 4.10 Latency profile

The recorded 500-image CURE configuration run had a mean elapsed time of 4.92 s per image (4,916.8 ms from the run summary, including startup and run overhead). The 235 gate-positive records had a mean elapsed time of 10.34 s and a median of 7.23 s; the 265 gate-negative records had a mean of 96.9 ms, falling to 25.7 ms over 264 records after excluding the first model-load record. Fig. S13 reports these observed strata and the raw-record all-image mean (4.92 s). A separate gate-only benchmark reported approximately 17 ms per image, but its 500-image evaluator elapsed time was 11.31 s including setup.

### 4.11 Inference-time architecture and deployment matrix

The full inference pipeline is shown in Fig. 2; the VRAM topology in Fig. S2. The evaluated configuration is the full dual-layer research prototype. A prior gate-only artifact is retained solely as historical context:

- **Historical gate-only output-routing benchmark (excluded from primary evidence).** The R32 artifact reported 76.2% binary output routing (198/250 NORMAL gate-negative and 183/250 ABNORMAL gate-positive), approximately 17 ms warm latency, and 0.35 GB VRAM. It used a historical CLS-token evaluator, whereas the production gate checkpoint declares masked mean pooling; it therefore cannot be interpreted as a measurement of the current gate implementation, the primary STRUCT endpoint, or clinical performance.
- **Full dual-layer research prototype.** VRAM 8.87 GB; observed mean elapsed time 4.92 s per image across the 500-image evaluation run and 10.34 s among gate-positive records; STRUCT 78.0%.
- **Exploratory high-score routing configuration (CURE + PG fallback, opt-in).** Output-presence sensitivity +5.6 pp and output-presence precision *−*5.6 pp, with STRUCT unchanged (paired exact McNemar *p* = 1.000 on the 500-image structural outcomes). Not enabled by default.

The R30-vs-R31 PG-fallback trade-off is shown in Fig. S14. The fallback changes output-presence rates in opposite directions while leaving the paired STRUCT result unchanged in this cohort; it is an opt-in exploratory configuration.

### 4.12 Gate score distribution

The gate score distribution on the 500-image VinDr-CXR evaluation is shown in Fig. S11 (evaluated gate output). The operating threshold is *τ* = 0.0557; the AUC was measured on the historical gate-development split. The observed 26.8% ABNORMAL gate-miss rate is a descriptive gate result, whereas the final CURE output-presence score is 78.0% after gate-positive NORMAL images can produce zero bboxes. Because split IDs were not preserved, the gate-development AUC is not an independent test estimate.

## 5 Discussion

### 5.1 STRUCT comparison without a ceiling claim

The principal finding is that the three primary configurations showed no statistically detectable STRUCT difference on this shared-pool VinDr-CXR cohort. The architecture therefore does not demonstrate a higher STRUCT score than the tested Lingshu prompt configurations. The NORMAL empty-output and ABNORMAL non-empty-output rates differed in opposite directions across systems. The MedGemma 1.5 SigLIP exploration is reported only as an unpaired exploratory direction because its full-pipeline records were not retained in the released artifact. The output-presence frontier is shown in Fig. 5.

### 5.2 Why prompt tuning is fragile

The 50-image pilot showed a 7-percentage-point STRUCT difference between the Lingshu prompt variants. On the 500-image shared-pool evaluation, the same comparison yielded *p* = 0.864; the pilot result is therefore exploratory rather than evidence of a reproducible prompt effect. Future prompt comparisons would benefit from a prespecified effect size and a held-out evaluation cohort.

### 5.3 Why SFT failed

We explored supervised fine-tuning of the CURE LoRA on VinDr-CXR (v4, v5) and observed two failure modes: (a) the v4 model collapsed to a generic “no finding” output despite the VinDr 70.7:29.3 NORM:ABN class ratio differing from the CURE pre-training distribution, and (b) the v5 model degraded the anatomical specificity of the report (Fig. S12). These observations are reported as development history; no supplementary artifact is claimed. We concluded that the SFT cost (14 GPU-hours per attempt, plus the cost of finding the right training data distribution) was not justified by the absence of a reproducible STRUCT improvement and the observed descriptive loss of anatomical specificity. The selected pos_weight = 6.0 was a training hyperparameter, not the empirical class ratio.

### 5.4 Limitations

STRUCT is a binary stratum-level output-presence proxy; it does not measure lesion-level localization, IoU, label correctness, report correctness, or clinical utility. This retrospective benchmark on VinDr-CXR was not evaluated for clinical workflow, triage, screening, or diagnosis. Historical gate split IDs were not preserved, so the cohort should not be interpreted as an independent test set. Any prospective clinical claim would require radiologist adjudication, prespecified effect sizes, and held-out evaluation on multi-institutional cohorts.

Additional limitations are the single-dataset evaluation, the single-language English reporting setting, and use of official VinDr-CXR annotations rather than consensus radiologist bboxes; inter-rater agreement for the official source annotations was not reported in the original release. Generalization to NIH ChestX-ray14, PadChest, MIMIC-CXR, and OpenCXR remains unverified.

### 5.5 Future work

Three directions follow directly from this study. First, multi-dataset evaluation on NIH ChestX-ray14, PadChest, and MIMIC-CXR, with the McNemar protocol replicated on each. Second, a multi-language reporting layer that decouples the language model from the CXR-grounding model. Third, a future prospective or retrospective study with prespecified radiologist adjudication could assess diagnostic accuracy, localization agreement, calibration, failure handling, and clinical workflow effects. Such a study would be needed to support any future triage, screening, confirmation, or clinical decision-support claim; this preprint makes none of those claims.

## 6 Conclusion

The dual-layer Gate-Before-Generate architecture CXRxVLM v2 yielded STRUCT 78.0% [Wilson 95% CI 74.2– 81.4] on the selected 500-image shared-pool VinDr-CXR cohort, with zero additional task-specific VLM training in this study and 769 trainable gate parameters. The three primary configurations showed no statistically detectable STRUCT difference in this retrospective benchmark. CURE had a higher ABNORMAL non-empty-output rate, whereas Lingshu had higher NORMAL empty-output rates.

## Data Availability

VinDr-CXR is available through PhysioNet under the provider’s access, data-use, and redistribution terms: https://physionet.org/content/vindr-cxr/1.0.0/. The source images are not redistributed. The canonical 500-image image-ID list, per-image JSONL outputs, gate checkpoint metadata, and supporting figures are associated with the project archive at Zenodo (DOI 10.5281/zenodo.21765487); access to the source images remains subject to the dataset provider’s terms. The present study used only the VinDr-CXR data and artifacts identified in this manuscript; MIMIC-CXR and PadChest-GR were not used for the reported evaluation. Historical gate split IDs were not preserved, so the archive does not support retrospective proof of train/evaluation disjointness.

## Code Availability

The repository contains analysis and evaluation code. Model weights, adapters, source images, and any restricted metadata are not redistributed unless permitted by their respective providers’ licenses, access agreements, and model terms. Users must obtain and comply with the applicable terms for VinDr-CXR, Rad-DINO, MedGemma, CURE, and Lingshu independently. Released image IDs, derived outputs, and figures should not be used to reconstruct or identify individuals.

## Ethics Statement

This retrospective analysis used VinDr-CXR data accessed through PhysioNet. The dataset is described by its provider as de-identified and subject to the provider’s data-use terms. No institution-specific determination about human-subjects research status, IRB approval, exemption, or waiver is claimed in this manuscript.

## Competing Interests

The authors declare no competing interests.

## Funding

This work was supported in part by personal equipment (an NVIDIA RTX 5090 workstation) and computing access provided by the Department of Computer Science and Engineering, National Taiwan Ocean University, and the Department of Applied Artificial Intelligence, Ming Chuan University. No specific grant funded this study.

## Author Contributions (CRediT)

**Tung-Chu Bai**: Conceptualization, Methodology, Software, Investigation, Data Curation, Formal Analysis, Visualization, Writing — Original Draft, Writing — Review & Editing. **Sheng-Cheng Yeh**: Supervision, Resources, Writing — Review & Editing.

## AI Tool Disclosure

The authors used large language models (Claude, GPT-4, and Microsoft Copilot) as drafting and verification aids during the preparation of this manuscript. Specifically, LLMs were used to (a) rephrase technical paragraphs for English-language fluency, (b) cross-check the consistency of statistical claims and confidence intervals against the underlying per-image JSONL records, (c) suggest related-work references, and (d) generate the matplotlib code for the supporting figures. All factual claims, statistical results, figure contents, and citations were independently verified by the authors against the underlying data, code, and primary references; the authors accept full responsibility for the content of the manuscript. No LLM was used to fabricate results, invent references, or replace authorial judgement.

## Data Availability

VinDr-CXR is available through PhysioNet under the provider's access, data-use, and redistribution terms: https://physionet.org/content/vindr-cxr/1.0.0/. The source images are not redistributed. The canonical 500-image image-ID list, per-image JSONL outputs, gate checkpoint metadata, and supporting figures are associated with the project archive at Zenodo (DOI https://doi.org/10.5281/zenodo.21765487); access to the source images remains subject to the dataset provider's terms. The present study used only the VinDr-CXR data and artifacts identified in this manuscript; MIMIC-CXR and PadChest-GR were not used for the reported evaluation. Historical gate split IDs were not preserved, so the archive does not support retrospective proof of train/evaluation disjointness.

## Supplementary Materials

**Figure S1.**
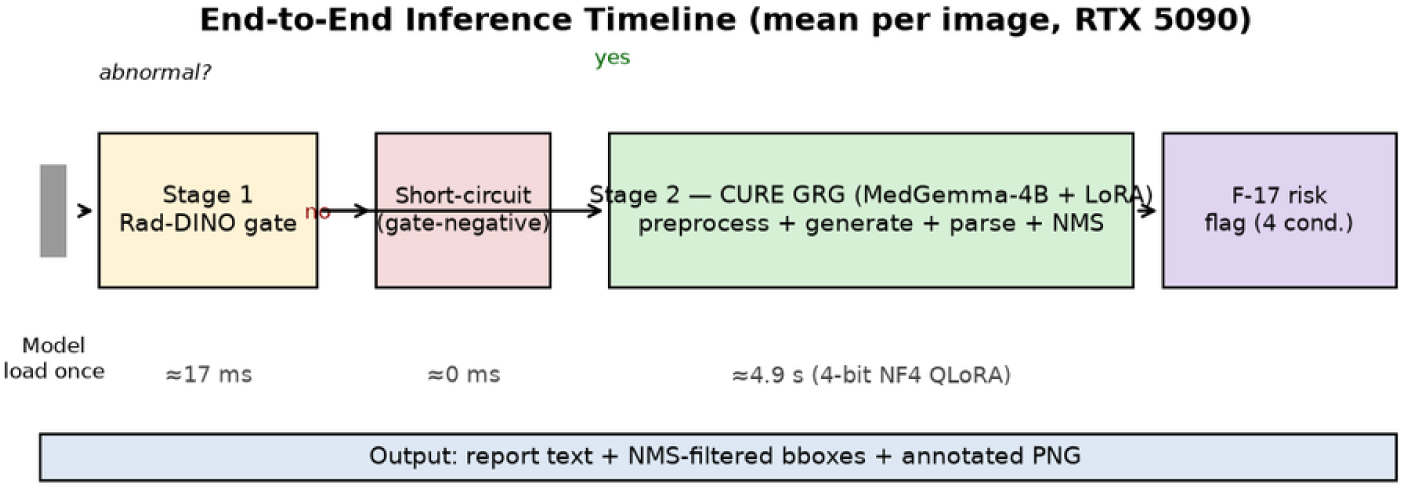
Inference timeline for the gate-only and full dual-layer paths.

**Figure S2.**
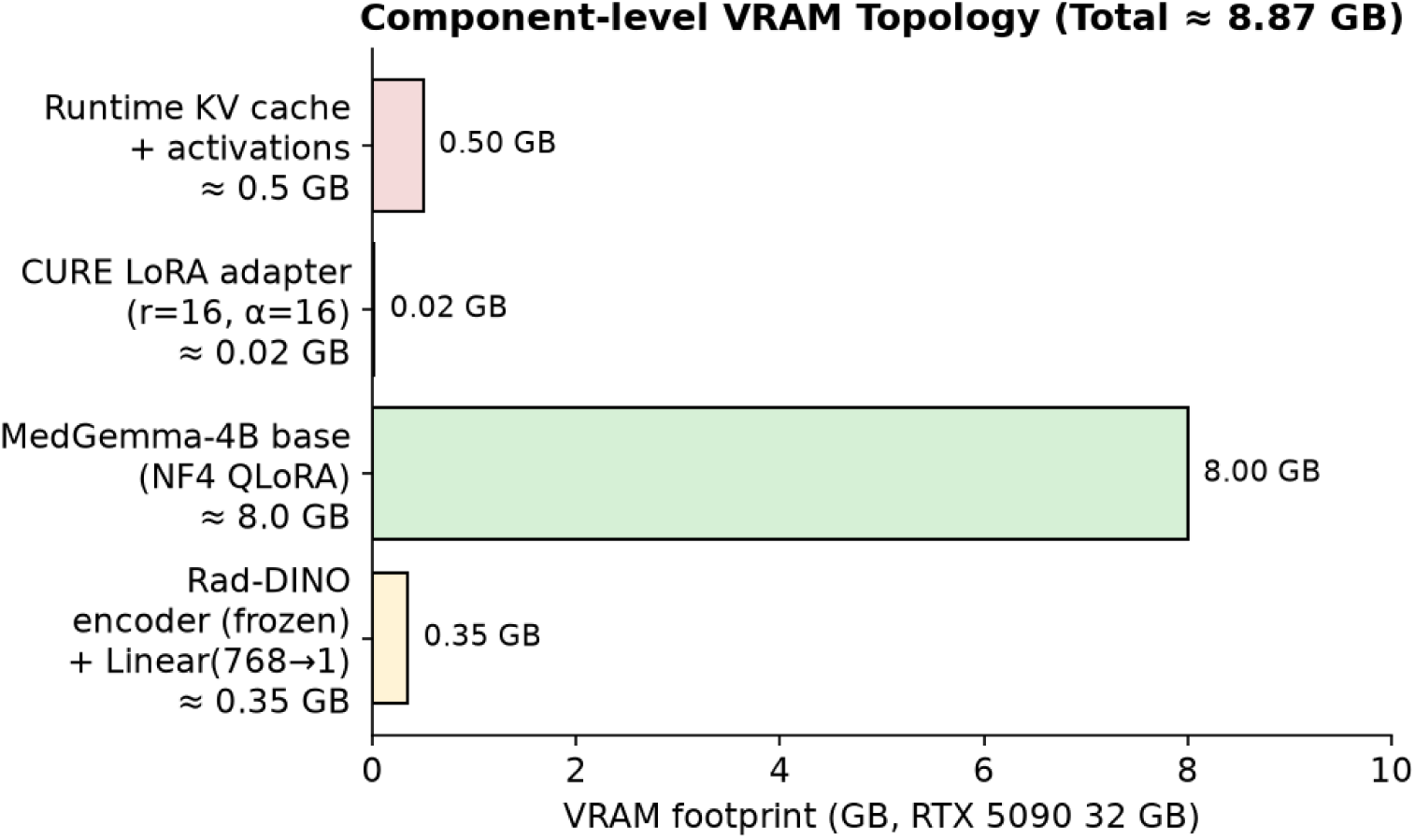
Component-level VRAM topology.

**Figure S3.**
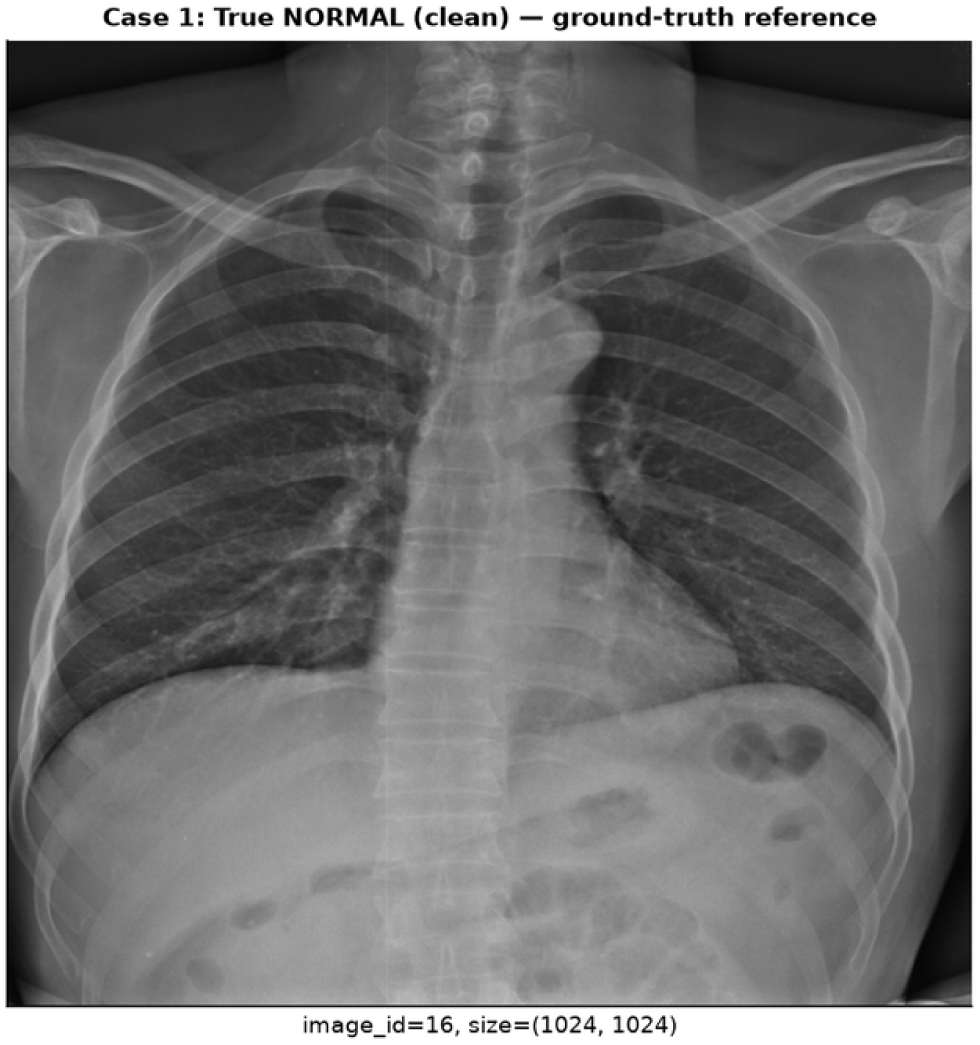
True NORMAL clean — ground-truth reference (no model prediction overlay).

**Figure S4.**
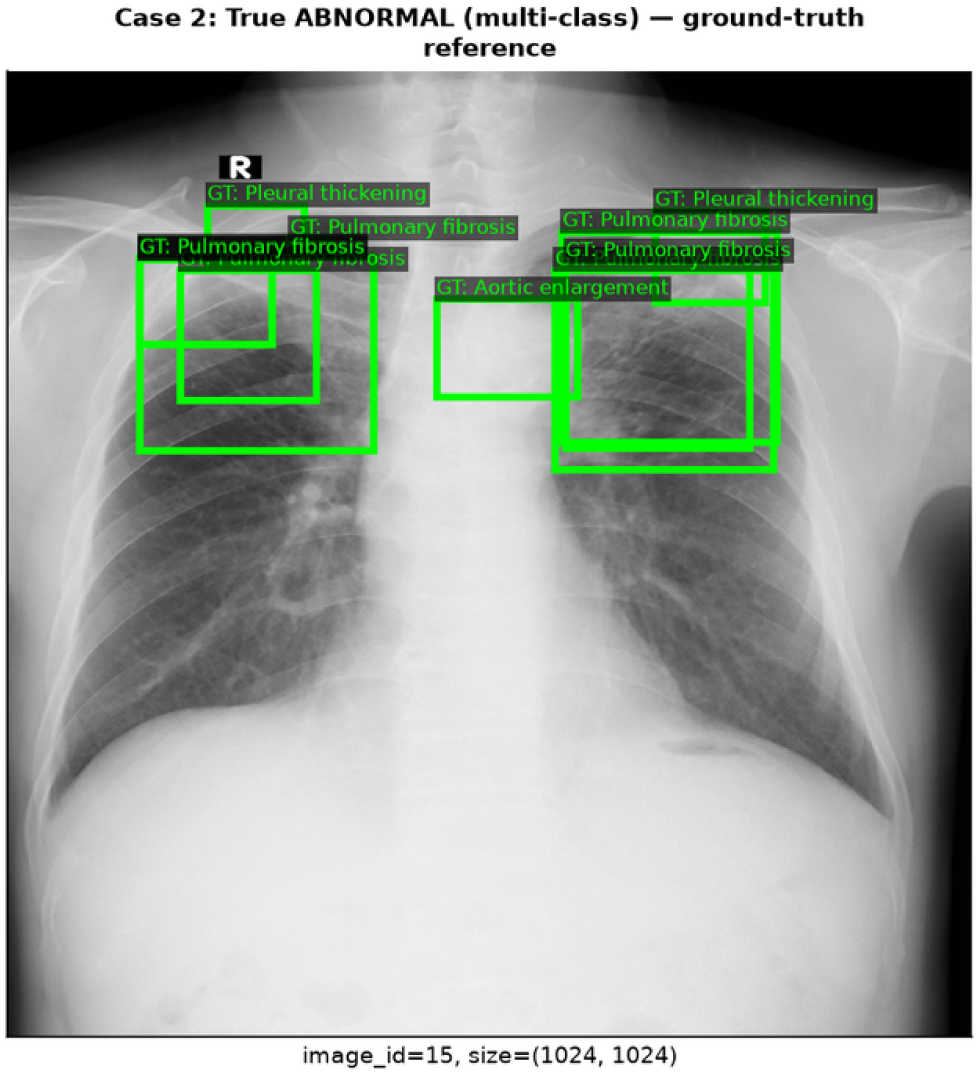
True ABNORMAL with multi-class findings — ground-truth reference (no model prediction overlay).

**Figure S5.**
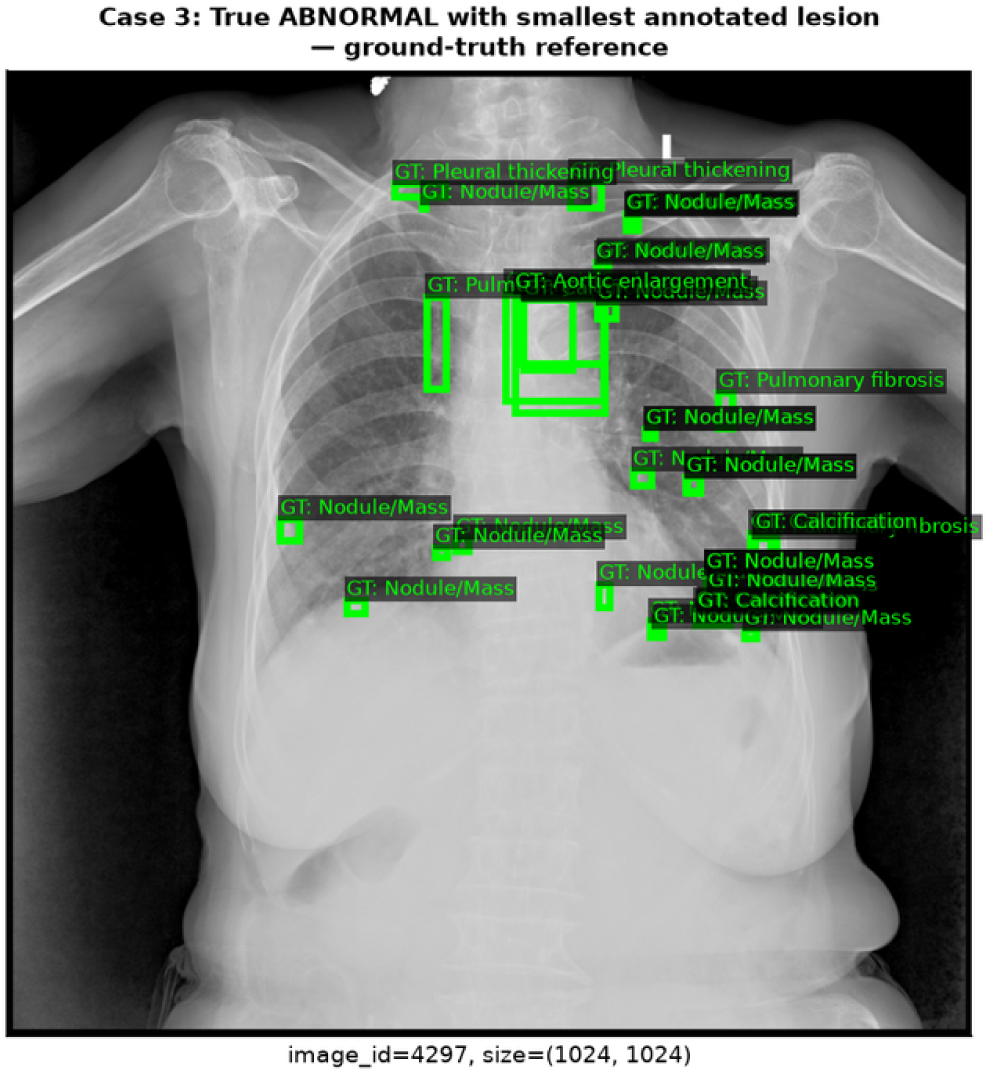
True ABNORMAL with the smallest annotated lesion in the source set — ground-truth reference (no model prediction overlay).

**Figure S6.**
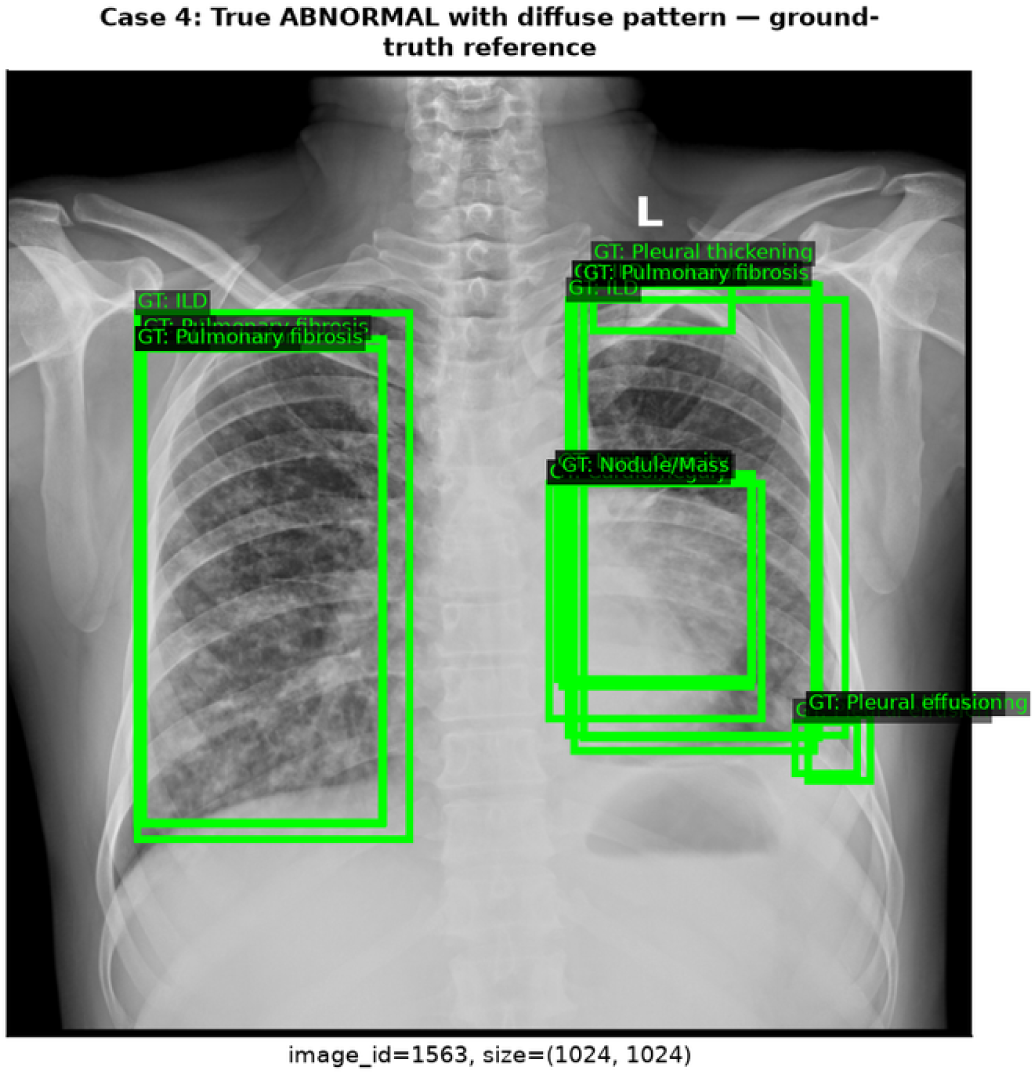
True ABNORMAL diffuse pattern — ground-truth reference (no model prediction overlay).

**Figure S7.**
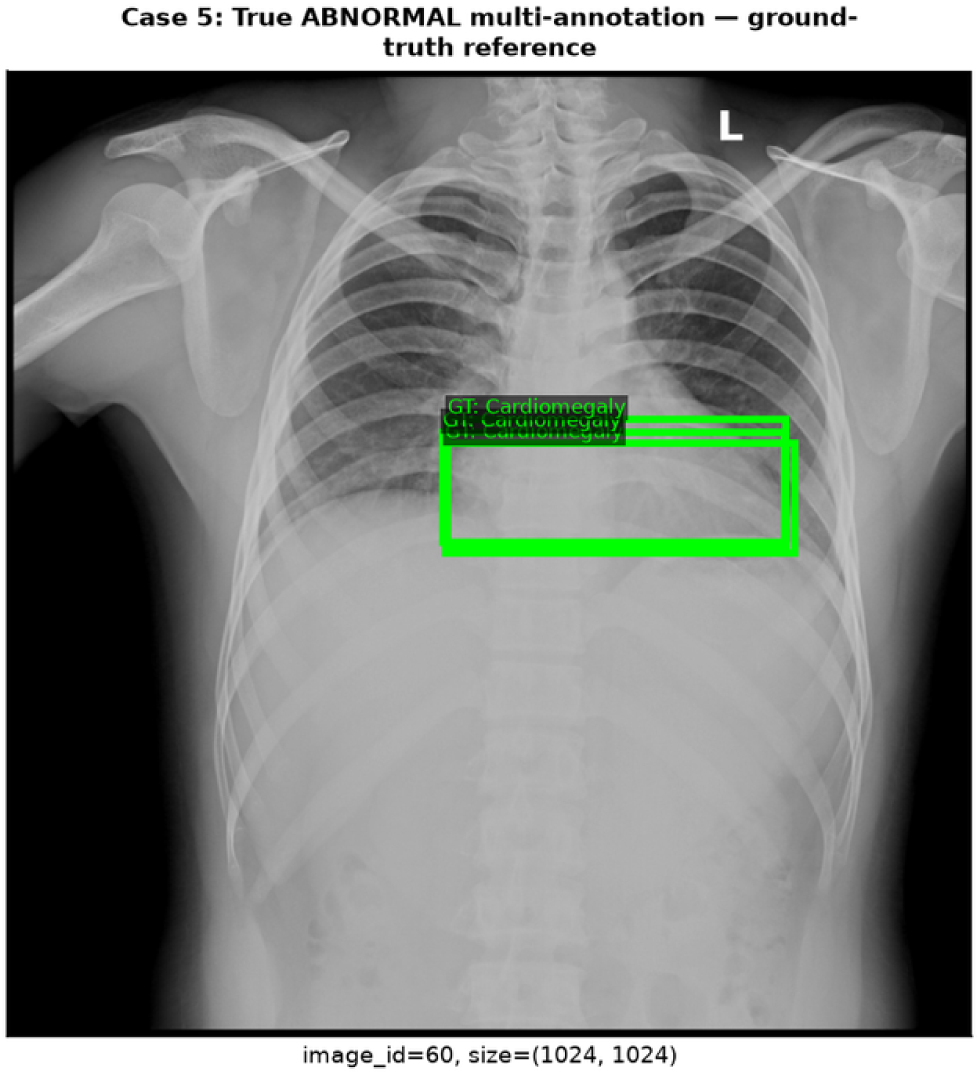
True ABNORMAL multi-annotation pattern — ground-truth reference (no model prediction overlay).

**Figure S8.**
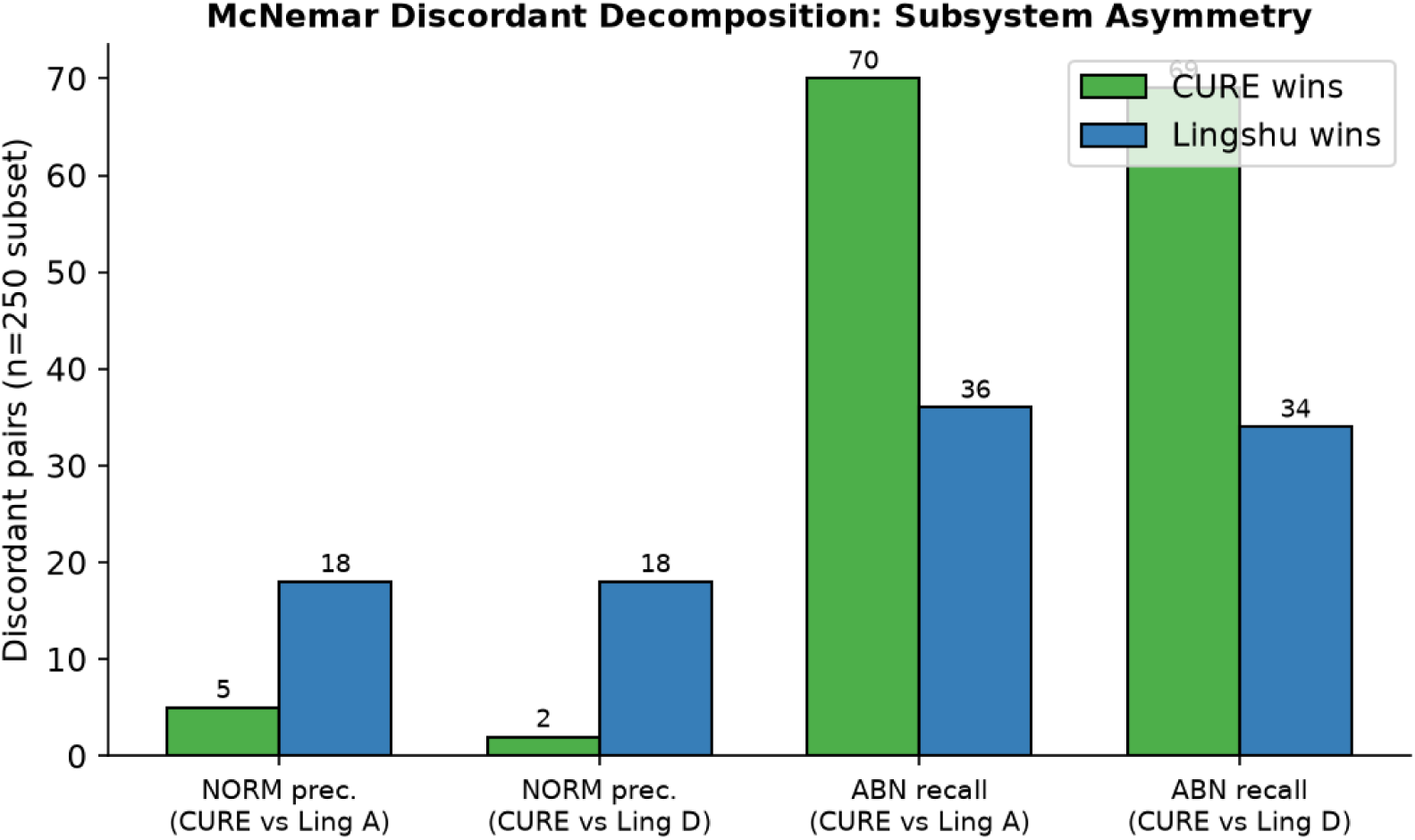
McNemar discordant-pair decomposition.

**Figure S9.**
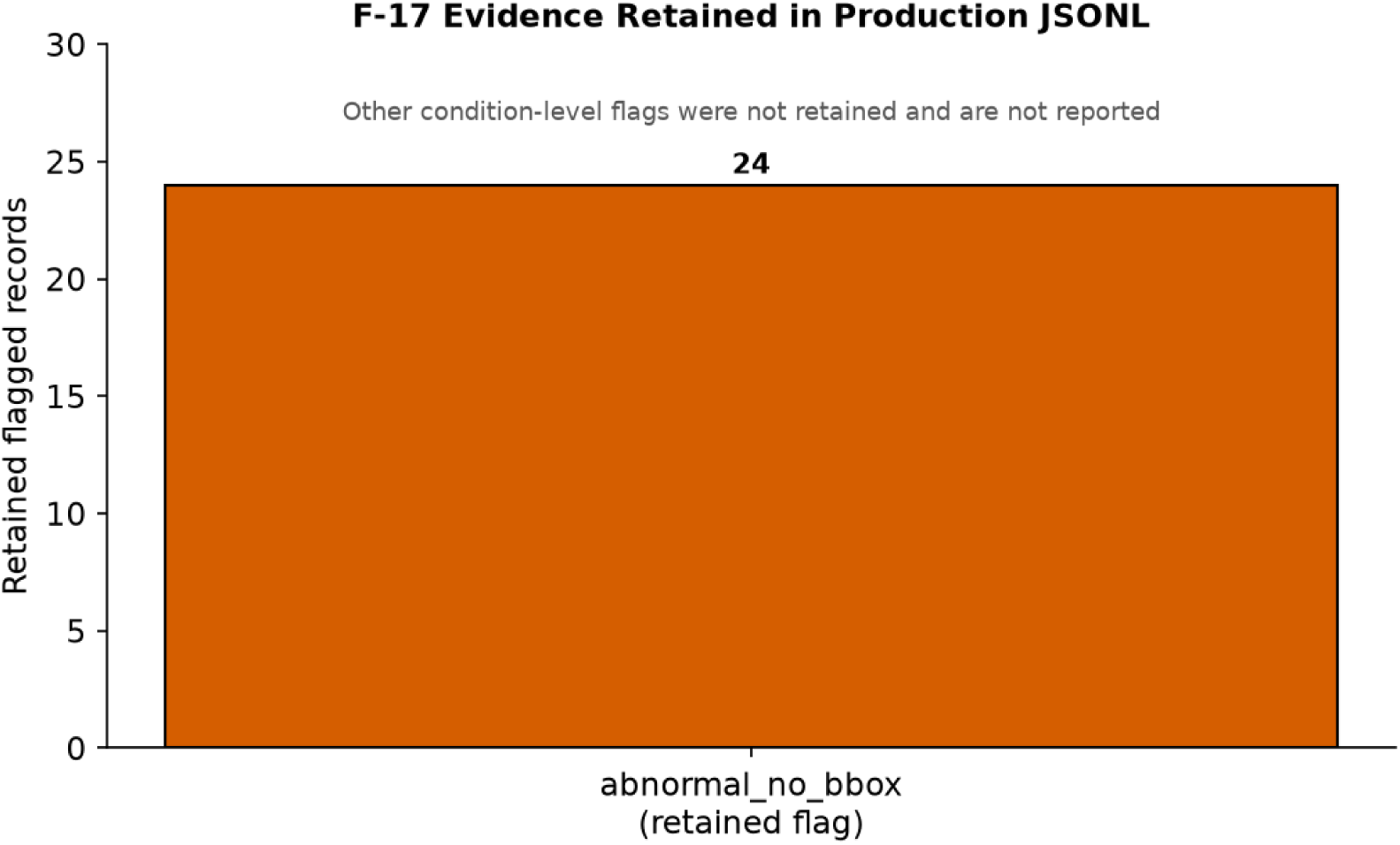
Retained F-17 evidence in the gate-positive ground-truth ABNORMAL audit (*n* = 183).

**Figure S10.**
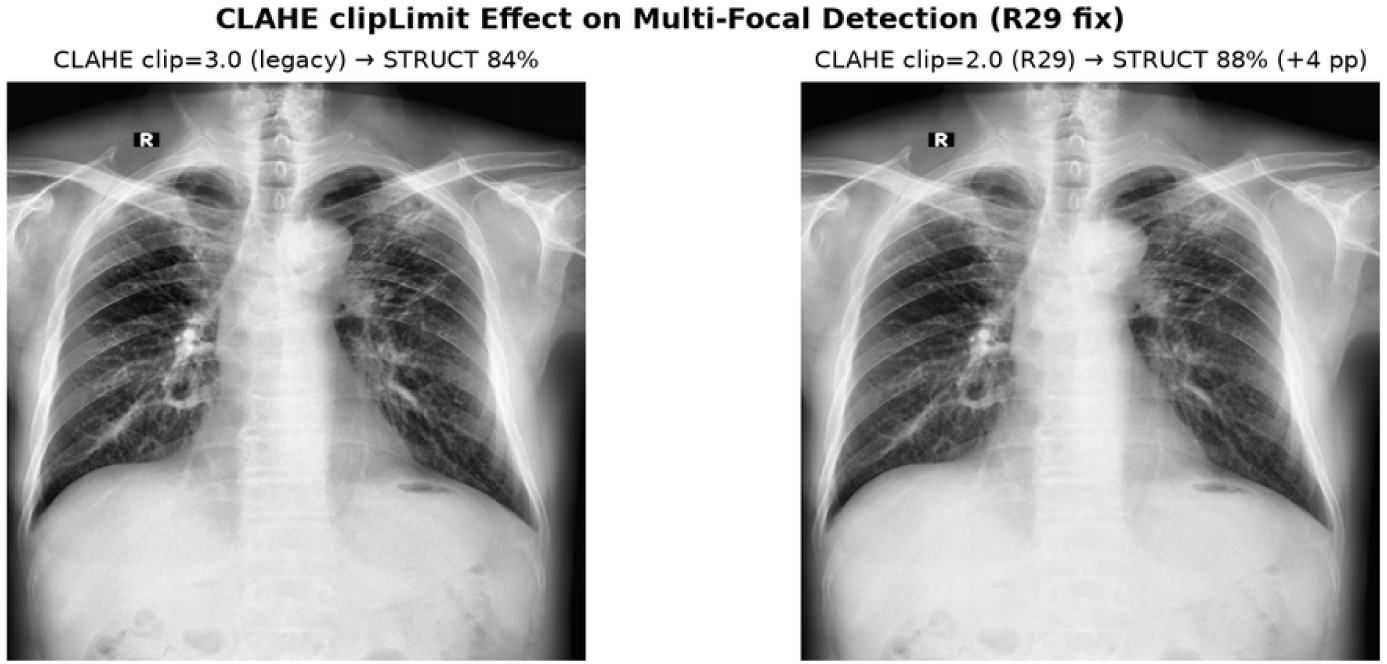
CLAHE clipLimit ablation.

**Figure S11.**
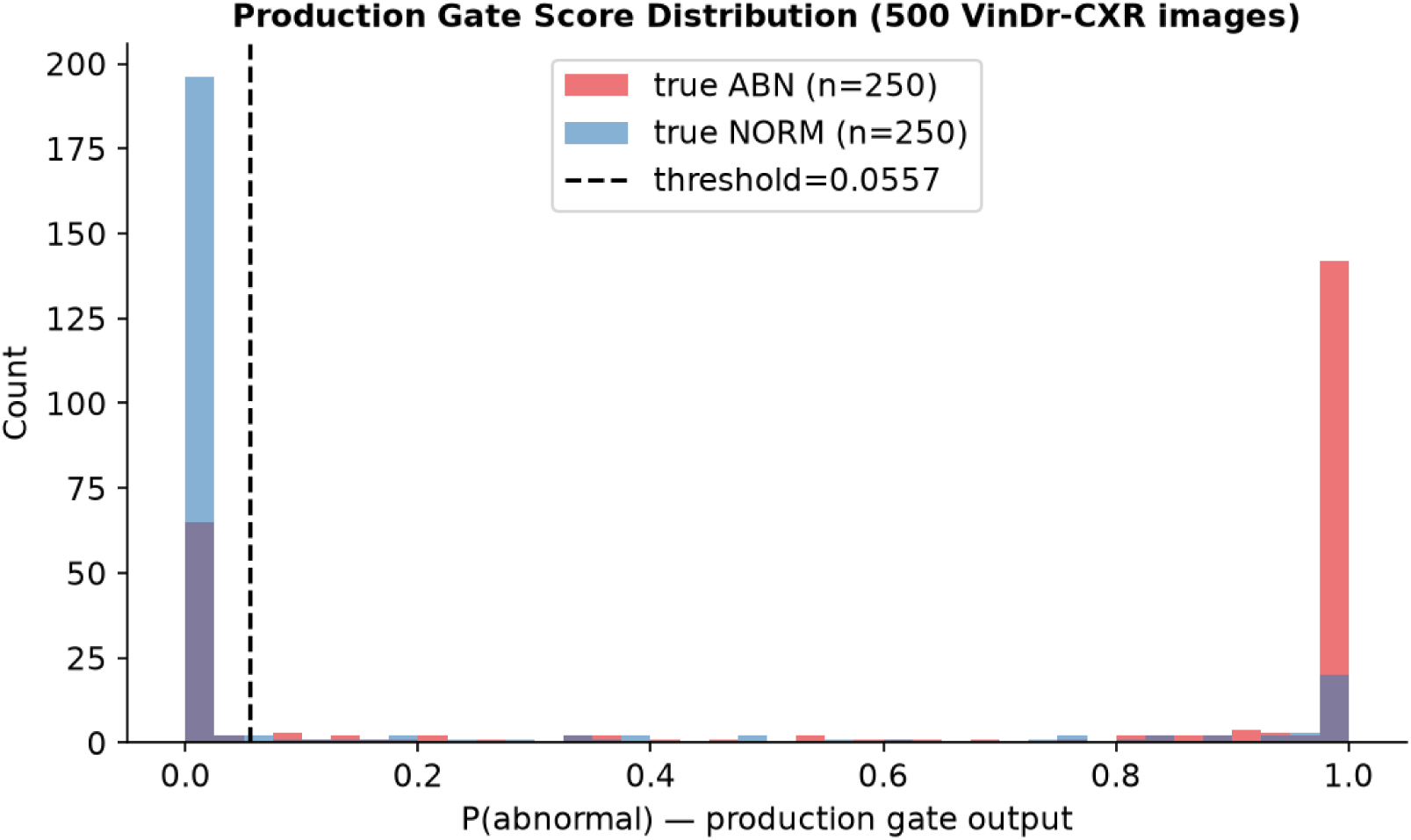
Gate score distribution on the 500-image VinDr-CXR evaluation (evaluated gate output).

**Figure S12.**
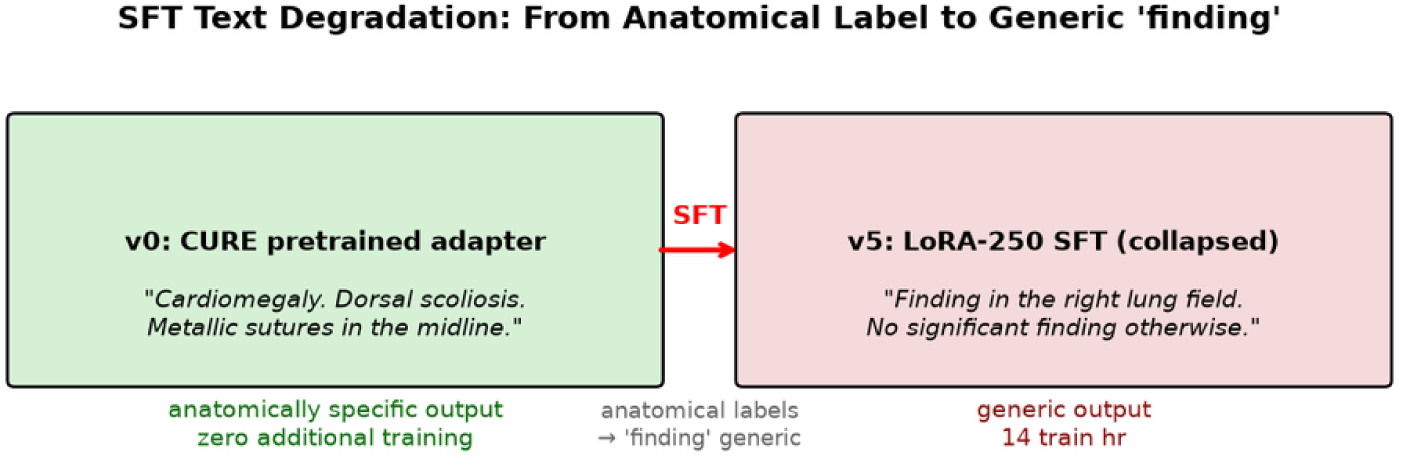
SFT text-quality degradation (pretrained CURE adapter vs LoRA-250 SFT).

**Figure S13.**
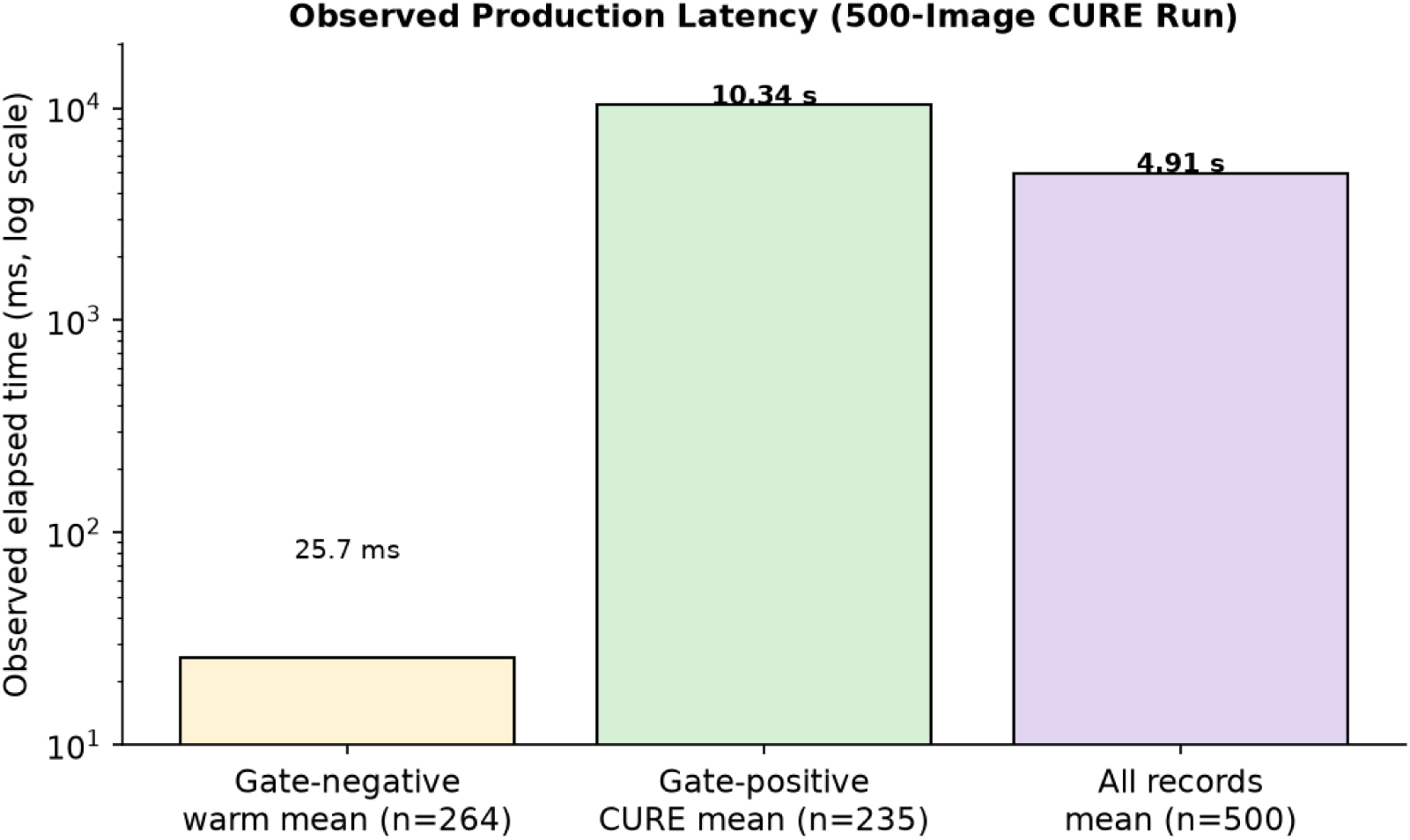
Observed configuration latency means from the 500-image CURE run.

**Figure S14.**
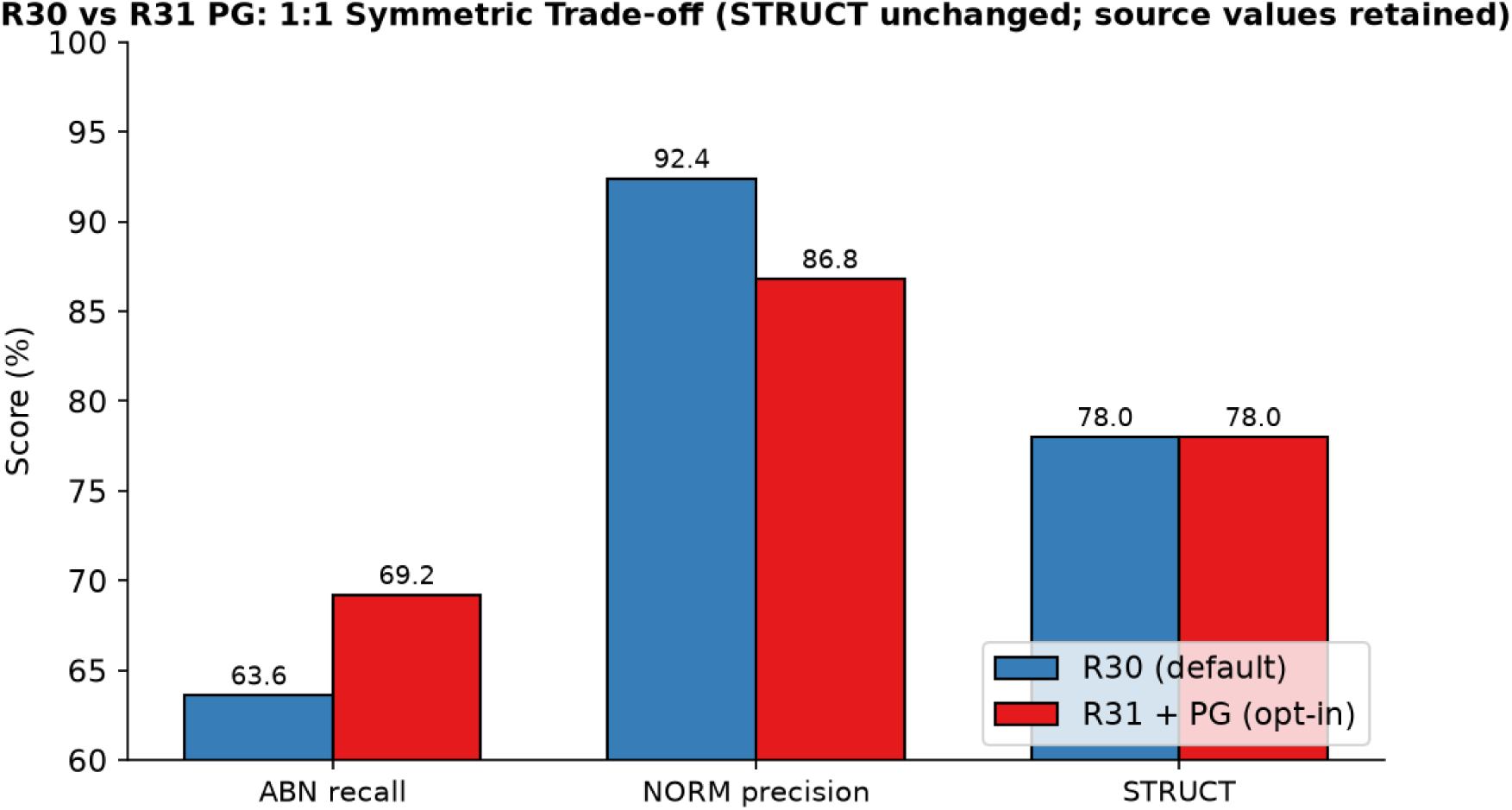
R30 vs R31 PG fallback (1:1 symmetric trade-off).

## Notes

### Competing Interest Statement

The authors have declared no competing interest.

### Author Declarations

VinDr-CXR: An open dataset of chest X-rays with radiologist's annotations was published in Scientific Data (2022) and has been publicly available through PhysioNet since its release. The dataset can be accessed at: https://physionet.org/content/vindr-cxr/1.0.0/ The dataset was openly available before the initiation of this study and requires no special access request, IRB approval, or ethics committee determination for use of the de-identified images and annotations. All 500 images used in this evaluation were randomly sampled with seed=42 from the official VinDr-CXR train.csv pool. Supporting analysis code and derived outputs (500-image image-ID list, per-image JSONL predictions, gate checkpoint metadata, and supplementary figures) are archived at Zenodo: https://doi.org/10.5281/zenodo.21765487

